# A Multi-Institutional Study Benchmarking Cycle Threshold Values for Major Clinical SARS-CoV-2 RT-PCR Assays

**DOI:** 10.1101/2022.06.22.22276072

**Authors:** J.E. Kirby, A. Cheng, M.H. Cleveland, E. Degli-Angeli, C.T. DeMarco, M. Faron, T. Gallagher, R.K. Garlick, E. Goecker, R.W. Coombs, C. Huang, R. Louzao, T.N. Denny, E. Morreale, G. Oakley, G. Reymann, A. Schade, S. Scianna, G.J. Tsongalis, P.M. Vallone, J. Huggett, N.A. Ledeboer, J.A. Lefferts

## Abstract

Real-time, reverse transcriptase PCR assays are a pervasive technology used for diagnosis of SARS-CoV-2 infection. These assays produce a cycle threshold value (Ct) corresponding to the first amplification cycle in which reliable amplification is detected. (1)Such Ct values have been used by clinicians and in public health settings to guide treatment, monitor disease progression, assess prognosis, and inform isolation practices. To understanding the risk of reporting out uncalibrated Ct values and potential for instead reporting out calibrated viral load values, we performed a multi-institutional study to benchmark major clinical platforms against a calibrated standard. We found that for any given Ct value, corresponding viral loads varied up to 1000-fold among the different tests. In contrast, when these different assays were calibrated against a common standard and then used to test unknown de-identified specimens at several dilutions, viral load values showed high precision between methods (standard deviation and range of 0.36 and 1.1 log10 genome copies) and high accuracy compared with droplet digital PCR (ddPCR) determinations (difference between mean CDC N2 and Sarbeco E ddPCR determinations and mean determinations by calibrated RT-PCR assays examined in our study of 0.044 log10 genome copies). We, therefore, find strong support for calibration of SARS-CoV-2 RT-PCR tests to allow conversion of cycle thresholds to accurate and precise viral load values that are reproducible across major clinical systems. Implementation of calibrated assays will provide more reliable information for clinical decision making and allow more rigorous interpretation of SARS-CoV-2 laboratory data in clinical and laboratory investigation.

---

Real-time, reverse transcriptase PCR assays are a pervasive technology used for diagnosis of SARS-CoV-2 infection. These assays are inherently quantitative. They produce a cycle threshold value (Ct) corresponding to the first amplification cycle in which reliable amplification is detected. The Ct in turn correlates with sample viral load, higher Ct values indicating a lower viral load and the converse. Despite current assays in the United States receiving Emergency Use Authorization as qualitative tests, the US Food and Drug Administration has uniquely allowed reporting of Ct values while simultaneously warning against their possible misinterpretation as a quantitative measure of viral load (1). The Ct values have been variously used by clinicians and in public health to guide treatment, monitor disease progression, assess prognosis, and inform isolation practices (2–9). However, Ct values are known to vary between platforms and assays for any given quantity of virus, therefore, generating debate about their use (8, 10). We have previously advocated that, as for quantitative viral load assays used in clinical practice, Ct values for SARS-CoV-2 RT-qPCR assays should be calibrated against a universal standard and reported out as platform agnostic, viral load values (i.e., genome copies/mL or IU/mL) (11).

To inform our understanding of the relationship of Ct values to viral loads and the potential risk of reporting out uncalibrated Ct values, we therefore initiated a multi-institutional study to benchmark major clinical platforms against a calibrated standard. The reference standard was a replication-incompetent, enveloped, positive-stranded RNA Sindbis virus (AccuPlex® Technology) into which the entire genome of SARS-CoV-2 is cloned. AccuPlex provides a safe, stable, biologically compelling surrogate for SARS-CoV-2, as it should behave similarly to live SARS-CoV-2 virus during each phase of RT-qPCR assay testing from extraction through detection. Ten-fold serial dilutions of this standard (prepared in viral transport media) were quantified independently by the US National Institute of Standards and Technology (NIST) and LGC NML by droplet digital PCR (ddPCR, CDC N2 (12) and Sarbeco E (13) primers and probes), with close agreement of genome copies/mL values, and distributed to participants.

Figure 1 shows the relationship of cycle threshold values to viral loads in genome copies/mL for platforms used at the participating institutions. Several points should be noted: (1) cycle threshold values from identical assays used in different laboratories were very consistent (i.e., Alinity and m2000); (2), the Abbott m2000 cycle threshold values are outliers as initial PCR cycles are not counted in its cycle threshold tally; (3) among the remaining assays, for any given viral load cycle threshold values varied over a range of ten cycles. Or stated another way, for any given cycle threshold value, corresponding viral loads (expressed as copies/mL) varied by 3.0 log10 (1000-fold) among the different tests. This is far greater than the ≤ 0.5 log10 (approximately three-fold) variance that is generally considered an acceptable difference between viral load assays (14).

**Figure 1.**
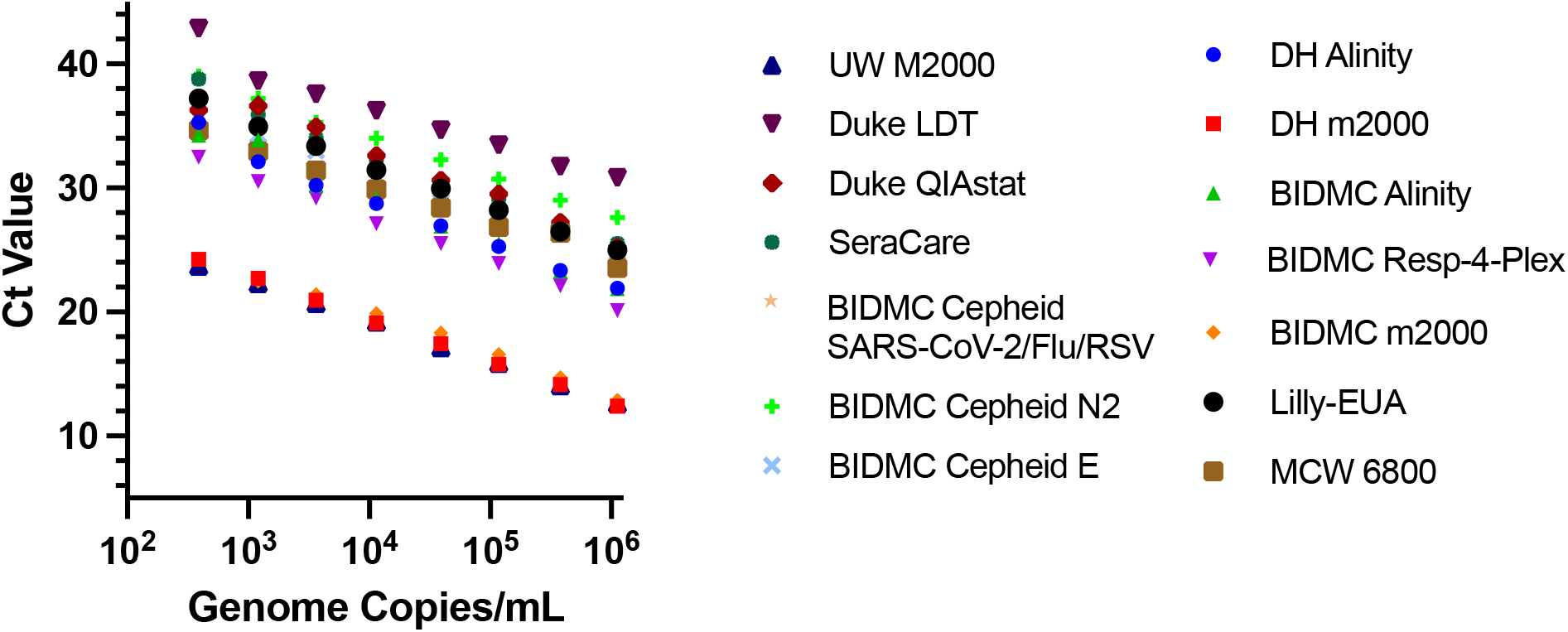
Cycle threshold values determined from calibrated standards. A series of dilutions of SARS-CoV-2 standards calibrated in genome copies/mL by droplet digital PCR were analyzed in a blinded fashion by different institutions on platforms in clinical use for SARS-CoV-2 testing. The legend includes site (abbreviations, BIDMC = Beth Israel Deaconess Medical Center, DH = Dartmouth Hitchcock Medical Center, UW = University of Washington Medical Center, EL = Eli Lilly, MCW = Medical College of Wisconsin, Duke = Duke University Medical Center, LGC NML = LGC National Measurement Laboratory, United Kingdom) and platform (abbreviations LDT = laboratory developed test; EUA = emergency use authorization; m2000 = Abbott RealTime SARS-CoV-2 assay; Alinity = Abbott Alinity m SARS-CoV-2 test; 6800 = cobas SARS-CoV-2 test run on cobas 6800; Resp-4-Plex = Abbott Alinity m Resp-4-Plex assay; Cepheid N2 and E = Ct values reported for separate targets from the Cepheid Xpress SARS-CoV-2 assay; Cepheid SARS-CoV-2/Flu/RSV = Cepheid Xpert Xpress SARS-CoV-2/Flu/RSV assay; and ddPCR = droplet digital PCR using CDC N2 (12) and Sarbeco E (13) primers and probes). Duke LDT used automated extraction and Sarbeco E primers and probe.

These data highlight compelling need for calibration of SARS-CoV-2 RT-qPCR assays against a universal standard and reporting out viral load values rather than potentially misleading Ct values. To demonstrate the utility of such an approach, we established assay-specific calibration curves from the data obtained in Figure 1. We then distributed serial dilutions of several de-identified patient specimens, procured from Serologix (Doylestown, PA) with ethical approval from the ADVARRA Internal Review Board (Columbia, MD, USA), to participating institutions. Ct values obtained were then converted to viral loads based on calibration data and compared to values obtained using ddPCR.

Notably, as shown in Figure 2, there was high precision among viral load results for calibrated RT-qPCR assays. Including all RT-qPCR data, the mean standard deviation and range of log10 genome copies for determinations were 0.36 and 1.1, respectively. Excluding the Duke QIAstat, Duke LDT, and Lilly EUA, assays, the average standard deviation and range were 0.2 and 0.65, respectively. The difference between mean ddPCR log10 genome copies/mL determinations for CDC N2 (12) and Sarbeco E (13) targets and mean log10 determinations for RT-qPCR methods was 0.044 log10 genome copies (0.077 log10 genome copies excluding QIAstat, Duke LDT, and Lilly EUA), indicating high accuracy of calibrated viral load results. The lower values obtained from QIAstat for specific samples suggests either inhibition and/or lower assay precision.

**Figure 2.**
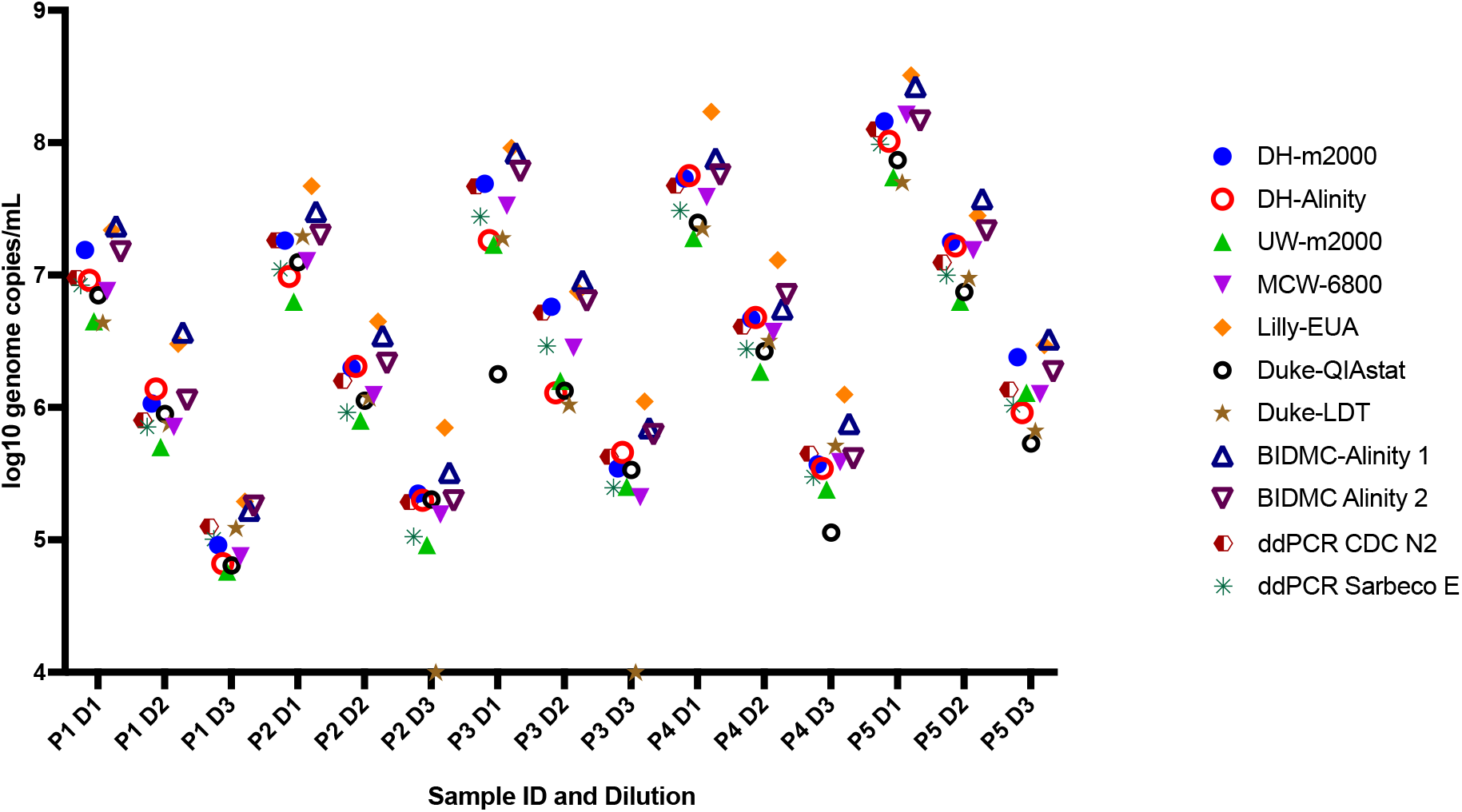
Determination of viral loads from patient samples based on calibration curves across platforms. Five patient samples (corresponding to P1 through P5) were serially diluted (corresponding to dilutions D1 through D3) and distributed to testing centers in a blinded fashion. Abbreviations for site of testing and assay are as indicated in the Figure 1 legend. BIDMC tested samples on two different Abbott Alinity m platforms listed here as 1 and 2. Droplet digital PCR was performed using a TaqMan digital PCR assay at LGC-SeraCare and LGC NML using CDC N2 and Sarbeco E primers and probes for amplification and detection. For ease of reference ddPCR results are shown on the left of each scatterplot for individual samples.

In summary, we demonstrated that Ct values determined on commonly used clinical platforms vary widely. In contrast, we find strong support for calibration of SARS-CoV-2 RT-qPCR tests against a recombinant standard to provide conversion of Ct values to viral load values that are accurate and reproducible across clinical systems. Implementation of calibrated assays will provide more reliable information for clinical decision making and allow more rigorous interpretation of SARS-CoV-2 laboratory data in clinical and laboratory investigations.

## Data Availability

All data produced in the present work are contained in the manuscript

## Funding/Acknowledgement

We thank LGC SeraCare (Gaithersburg, MD) for provision of SARS-CoV-2 AccuPlex calibration material and human specimens used in this study. J.E.K, A.C., E.D.A., E.G., and R.C. have received SARS-CoV-2 reagents from Abbott Molecular under a under COVID-19 Diagnostics Evaluation Agreements unrelated to this study. R.G., E.M., and C.H. are employees of LGC SeraCare. G.O. and A.S. are employees of Eli Lilly and Company and compensated with salary and stock. N.A.L. has received consulting honoraria from LGC. All other authors, no conflicts of interest. Certain commercial equipment, instruments, or materials are identified in this paper in order to specify the experimental procedure adequately. Such identification is not intended to imply recommendation or endorsement by the National Institute of Standards and Technology nor is it intended to imply that the materials or equipment identified are necessarily the best available for the purpose.

